# Clinical evaluation of the molecular-based BD SARS-CoV-2/Flu for the BD MAX™ System

**DOI:** 10.1101/2021.02.23.21251915

**Authors:** Sonia Paradis, Elizabeth Lockamy, Charles K. Cooper, Stephen Young

**Author notes:** **To whom correspondence should be addressed:** Stephen Young, PhD, Address: 1001 Woodward Place, N.E. Albuquerque, NM 87102. To whom correspondence should be addressed (alternate): Charles K. Cooper, MD, Vice President of Medical Affairs, Becton, Dickinson and Company, BD Life Sciences – Integrated Diagnostic Solutions 7 Loveton Circle, Sparks MD 21152, USA.

## Abstract

**Objectives:** Establish the diagnostic performance related to SARS-CoV-2 and Flu A/B detection for the BD SARS-CoV-2/Flu for BD MAX™ System (“MAX SARS-CoV-2/Flu”) multiplex assay.

**Methods and Materials:** Two hundred and thirty-five (235) retrospective nasopharyngeal specimens were obtained from external vendors. The BD BioGx SARS-CoV-2 Reagents for BD MAX™ System (“BioGx SARS-CoV-2”) and the Cepheid Xpert^®^ Xpress Flu/RSV (“Xpert Flu”) were utilized as reference methods.

**Results:** By reference methods, 52 specimens were SARS-CoV-2-positive, 59 were Flu A-positive, and 60 were Flu B-positive. MAX SARS-CoV-2/Flu had positive percent agreement (PPA) and negative percent agreement (NPA) values for SARS-CoV-2 detection of 96.2% ([95%CI]:87.0-98.9) and 100% [95%CI:88.7-100], respectively; PPA values for Flu A and Flu B of 100% [95%CI:93.9-100] and 98.3% [95%CI:91.1-99.7], respectively, and NPA values for Flu A and Flu B of 98.9% [95%CI:94.0-99.8] and 100% [95%CI:95.9-100], respectively.

**Discussion:** The MAX SARS-CoV-2/Flu assays met FDA-EUA performance criteria for SARS-CoV-2 (≥95% for PPA and NPA) and FDA clearance criteria for Flu A/B (PPA ≥90%; lower bound of the 95%CI ≥80%) and (NPA ≥95%; lower bound of the 95%CI ≥90%).

## INTRODUCTION

Since the report of the first cluster of COVID-19 cases in December 2019, over 147 million COVID-19 cases and 3 million COVID-19-related deaths worldwide have been reported by the end of April, 2021, and the numbers continue to rise.(1) In the US, more than 32.1 million COVID-19 cases and over 572,000 COVID-19 deaths have been recorded through end of April 2021.(2) Although it seems that the 2020-2021 influenza season will not impact the health care systems, the 2019-2020 flu season resulted in over 38 million cases involving symptomatic illness and approximately 22,000 deaths in the US.(3) Each year, there are an estimated 1 billion cases of influenza globally, of which, 3-5 million are severe cases and 29,000-655,000 lead to influenza-related respiratory deaths.(4) Although as of April 2021 influenza activity is low,(5) this virus has the potential of increasing the workload of healthcare workers already overwhelmed by COVID-19.

While COVID-19 and influenza spread through a similar mechanism of transmission and have overlapping symptoms, including cough and fever, the isolation length and the therapeutic approach for COVID-19 patients and influenza patients are not uniform.(6) The recommended isolation period after symptoms onset is a minimum of 4-5 days for flu,(7) whereas it is a minimum of 10 days for COVID-19,(8) impacting absenteeism and contact tracing. Additionally, the therapeutic interventions vary between these two diseases. The impact of anti-viral drug therapy, such as Tamiflu^®^ or Xoflusa, which have been used for influenza patients,(9) are not approved or their efficacy well-understood for patients with COVID-19. Therefore, safety concerns may preclude any potential efficacy. A similar concern exists for drugs such as remdesivir and corticosteroids, which have been used to treat COVID-19 patients, but are not approved for use in influenza patients, and may (for example, in the case of corticosteroids) have negative side effects in influenza patients.(6) As the society gradually reopens the social interactions in many places, the potential coincidence of both COVID-19 and influenza cases in high numbers during respiratory virus seasons is a significant concern. Especially since the respiratory virus activity usually peaks between December and February in the US,(10) and in future respiratory virus seasons could produce a significant strain on the healthcare system. Therefore, differential diagnosis of COVID-19 and influenza, will be an important component for proper patient triage, management, and treatment.

Molecular diagnostics for the detection of SARS-CoV-2, including real-time polymerase chain reaction (RT-PCR) assays, have played an important role in the detection of SARS-CoV-2 and diagnosis of COVID-19 due to their high sensitivity.(11) Similarly, RT-PCR-based detection of Influenza A/B (“Flu A/B”) virus nucleic acid has been established for a number of years and is commonly employed to establish an influenza diagnosis.(12) Due to the expected co-circulation with the potential co-infection of SARS-CoV-2 and Flu A/B, a multiplex RT-PCR assay, for detection all three targets, could help provide faster results and improve patient management and treatment.(13) This report describes the performance of the new BD SARS-CoV-2/Flu assay reagents for BD MAX™ System multiplex assay for detection of SARS-CoV-2 and Flu A/B. The comparator reference methods were the BD BioGx SARS-CoV-2 Reagents for BD MAX™ System and Cepheid Xpert^®^ Xpress Flu/RSV. The objective here was to determine the performance characteristics of the new multiplex BD SARS-CoV-2/Flu assay.

## MATERIALS AND METHODS

### Specimens and assays

This study, which was conducted as part of a Food and Drug Administration-Emergency Use Authorization (EUA) submission, included data comparing the BD SARS-CoV-2/Flu for BD MAX™ System (“MAX SARS-CoV-2/Flu;” Becton, Dickinson and Company; BD Life Sciences—Integrated Diagnostics Solutions, Sparks, MD, USA) with reference methods, BD BioGx SARS-CoV-2 Reagents for BD MAX™ System (“BioGx SARS-CoV-2;” Becton, Dickinson and Company; BD Life Sciences – Integrated Diagnostics Solutions, Sparks, MD, USA) and Cepheid Xpert^®^ Xpress Flu/RSV (“Xpert Flu;” Cepheid^®^, Sunnyvale, CA, USA), for detection of SARS-CoV-2 and Flu A/B, respectively.(14) The BioFire^®^ Respiratory 2.1 Panel (“BioFire SARS-CoV-2;” BioFire Diagnostics, Salt Lake City, UT, USA) was used to test specimens for which MAX SARS-CoV-2/Flu and BioGx SARS-CoV-2 provided discrepant results; the cobas^®^ Influenza A/B & RSV assay for use on the cobas^®^ Liat^®^ System (“Liat Flu;” Roche Diagnostics, Indianapolis, IN, USA) was used to test specimens for which MAX SARS-CoV-2/Flu and Xpert Flu provided discrepant results. All assays were performed according to each manufacturer’s instructions for use. MAX SARS-CoV-2/Flu, BioGx SARS-CoV-2, and BioFire SARS-CoV-2 assays were performed at BD Integrated Diagnostics Solutions; Xpert Flu and Liat Flu assays were performed at TriCore Reference Laboratories (Table S1).

Nasopharyngeal specimens for SARS-CoV-2 testing were obtained from New York Biologics, Inc. (Southhampton, NY, USA) and Trans-Hit Bio (Laval, QC, Canada), and nasopharyngeal specimens for Flu A/B testing were obtained from New York Biologics, Inc. Specimens provided by New York Biologics were collected under protocols approved by Western Institutional Review Board (WIRB) and from Ethical & Independent Review Services (EIRS). These institutional review board approvals provide a waiver of informed consent on collection protocols for de-linked and de-identified specimen collections. Samples obtained from Trans-Hit Bio were collected under their biobank umbrella protocol (approved by the Valleywise Health Institutional Review Board) that allows for the collection of various bio-specimens. Two hundred and thirty-five (235) nasopharyngeal specimens either in Copan Universal Transport Medium (UTM^®^) or in BD Universal Viral Transport (UVT) system were collected between November 30, 2019 to September 3, 2020. The samples were obtained from individuals with ages ranging from ≤5 years of age to ≥60 years of age and the residual transport media was stored at -65° C ∼ -80°C (Table 1).

**Table 1.** SARS-CoV-2 and influenza positivity by reference method or MAX SARS-CoV-2/Flu across age groups.

| Age group | Reference |  |  | BD MAX SARS-CoV-2/Flu |  |  |
| --- | --- | --- | --- | --- | --- | --- |
|  | SARS-CoV-2<br>n (%) | Influenza A<br>n (%) | Influenza B<br>n (%) | SARS-CoV-2<br>n (%) | Influenza A<br>n (%) | Influenza B<br>n (%) |
| **≤5 years** 21.6%<br>(n=50) | 0 (0.0) | 19 (32.2) | 13 (21.7) | 0 (0.0) | 19 (31.7) | 13 (22.0) |
| **6-21 years** 19.8%<br>(n=46) | 0 (0.0) | 12 (20.3) | 26 (43.3) | 0 (0.0) | 12 (20.0) | 26 (44.1) |
| **22-59 years**<br>41.8% (n=97) | 39 (75.0) | 16 (27.1) | 16 (26.7) | 37 (74.0) | 17 (28.3) | 15 (25.4) |
| **≥60 years** 16.8%<br>(39) | 13 (25.0) | 12 (20.3) | 5 (8.3) | 13 (26.0) | 12 (20.0) | 5 (8.5) |
| **Overall (N=232)<sup>a</sup>** | 52 | 59 | 60 | 50 | 60 | 59 |

<sup>a</sup>Compliant and reportable for MAX and comparator assays.

The specimens were collected as part of standard of care (SOC) and residual de-identified samples were frozen and used for this research. This article was prepared according to STARD guidelines for diagnostic accuracy studies reporting.(15)

### Data analysis

The primary outcome measures for this study were positive and negative percent agreement (PPA and NPA, respectively) point estimates (with 95% confidence interval [95% CI] calculated using the Wilson score method) for the MAX SARS-CoV-2/Flu assay, compared to each respective reference assay. The McNemar test was used for 2×2 classification to test the difference between paired proportions. The calculated difference is that of marginal proportions ([total proportion of SARS-CoV-2, Flu A, or Flu B positives] – [total proportion of positives (for each of the three causes) by clinical diagnosis]). A p-value <0.05 was utilized to distinguish significant differences (note here that a p-value >0.05 indicates only that disagreement between the two diagnostics methods is random). The Cohen’s kappa coefficient was utilized to gauge the agreement between two raters (reference and test) to classify results into mutually exclusive categories. Κ=(P_o_ ^-P^e)/1-P_e_(<0, 0, and >0 indicating agreements worse than, no better or worse than, and better than that expected by chance). Acceptance criteria for the MAX SARS-CoV-2/Flu assay for US FDA-EUA authorization for SARS-CoV-2 was ≥95% for both PPA and NPA.(16) The PPA criteria for Flu A/B was ≥90% (lower bound of the 95%CI ≥80%) and the NPA criteria for Flu A/B was ≥95% (lower bound of the 95%CI ≥90%). Only compliant and reportable results for both MAX SARS-CoV-2/Flu and comparator assays were included in this analysis.

### Data Availability

Data will be made publicly available upon publication and upon request for peer review.

## RESULTS

235 specimens were included in this study, from which, three were excluded due to unreportable results from an instrumental failure. From the remaining 232 specimens, reference method testing for SARS-CoV-2 (BioGx SARS-CoV-2) and Flu A/B (Xpert Flu), resulted in 52 positive SARS-CoV-2 specimens, 59 positive Flu A specimens, and 60 positive Flu B specimens (Table 1). By reference methods, 30, 91, and 90 specimens were negative, respectively, for SARS-CoV-2, Flu A, and Flu B. Among all positive cases, the 22-59 years age group had the highest SARS-CoV-2 positivity, the ≤5 age group had the highest Flu A positivity, and the 6-21 years age group had the highest Flu B positivity.

MAX SARS-CoV-2/Flu results were compared to results from each respective reference method to determine PPA and NPA values. MAX SARS-CoV-2/Flu had PPA and NPA values of 96.2% [95%CI: 87.0, 98.9] and 100% [95%CI: 88.7, 100], respectively, for detection of SARS-CoV-2. For Flu A, MAX SARS-CoV-2/Flu had PPA and NPA values of 100% [95%CI: 93.9, 100] and 98.9% [95%CI: 94.0, 99.8], respectively. For Flu B, MAX SARS-CoV-2/Flu had PPA and NPA values of 98.3% [95%CI:91.1, 99.7] and 100% [95%CI: 95.9, 100], respectively (Table 2).

**Table 2.** Performance of the MAX SARS-CoV-2/Flu assay for detection of SARS-CoV-2, Flu A and Flu B compared to reference.

|  | **SARS-CoV2<sup>a,c</sup>** | **Flu A<sup>b,c</sup>** | **Flu B<sup>b,c</sup>** |
| --- | --- | --- | --- |
| **PPA** | 96.2% [87.0%, 98.9%] | 100% [93.9%, 100%] | 98.3% [91.1%, 99.7%] |
| **NPA** | 100% [88.7%, 100%] | 98.9% [94.0%, 99.8%] | 100% [95.9%, 100%] |
| **MAX (+) / Ref (+)** | 50 | 59 | 59 |
| **MAX (+) / Ref (-)** | 0 | 1 | 0 |
| **MAX (-) / Ref (+)** | 2 | 0 | 1 |
| **MAX (-) / Ref (-)** | 30 | 90 | 90 |
| **kappa** | 0.948 | 0.986 | 0.986 |

**Abbreviations:** PPA, positive percent agreement; NPA, negative percent agreement
<sup>a</sup>Reference method was the BioGx SARS-CoV-2 RT-PCR assay.
<sup>b</sup>Reference method was the Xpert Flu RT-PCR assay.
<sup>c</sup>A statistically significant difference (via McNemar's test on paired proportions was not observed for MAX SARS-CoV-2/Flu for detection of SARS-CoV-2 (-2.4 [95% CI: -5.8, 0.0]; p=0.500), Flu A (0.67 [95% CI: -0.64, 1.97]; p=1.000), or Flu B (-0.67 [95% CI: -1.97, 0.64]; p=1.000).

During discordant testing, the MAX SARS-CoV2/Flu assay was in agreement with the third assays (i.e. BioFire SARS-Cov-2 assay and Liat Flu assay) for both SARS-CoV-2 negative results and for the Flu A positive result by the MAX SARS-CoV-2/Flu assay. For Flu B, the Liat Flu assay agreed with the Xpert Flu assay negative result. However, all discrepant results were associated with high cycle threshold (Ct) values (ranging from 37.8 to 39.5). The MAX SARS-CoV-2 showed 100% PPA in specimens with reference method results associated with Ct values ≤30 (Table 3).

**Table 3.** Comparison of MAX SARS-CoV-2/Flu assay results with those from the BioGx SARS-CoV-2 and Xpert Flu assays, stratified by cycle threshold category.

| BioGx SARS-CoV-2 |  |  |
| --- | --- | --- |
| MAX SARS-CoV-2 | Positive (Ct ≤ 30) | Positive (Ct > 30) |
| Positive | 41 | 9 |
| Negative | 0 | 2<sup>a</sup> |
| Total | 41 | 11 |
| PPA (95% CI) | 100% (91.4% - 100%) | 81.8% (52.3% - 94.9%) |

| MAX Flu A | Xpert Flu A<sup>b</sup> |  |
| --- | --- | --- |
| MAX | Positive (Ct ≤ 30) | Positive (Ct > 30) |
| Positive | 48 | 11 |
| Negative | 0 | 0 |
| Total | 48 | 11 |
| PPA (95% CI) | 100% (92.6% - 100%) | 100% (74.1% - 100%) |

| MAX Flu B | Xpert Flu B |  |
| --- | --- | --- |
| MAX | Positive (Ct ≤ 30) | Positive (Ct > 30) |
| Positive | 48 | 11 |
| Negative | 0 | 1<sup>c</sup> |
| Total | 48 | 12 |
| PPA (95% CI) | 100% (92.6% - 100%) | 91.7% (64.6% - 98.5%) |

**Abbreviations:** Ct, PCR cycle threshold; PPA, positive percent agreement
<sup>a</sup>One specimen corresponded to a Ct value for the N1 result = 39.5 and an N2 result = negative. One specimen corresponded to a N1 result = negative and a Ct value for the N2 result = 38.8. Discrepancy testing with the BioFire SARS-CoV-2 assay was positive (agreement with MAX) for both specimens.
<sup>b</sup>One specimen (not shown here) was positive by MAX (Ct value = 38.8) and negative by Xpert Flu. Discrepancy testing with the Liat Flu assay was positive (agreement with MAX SARS-CoV-2/Flu).
<sup>c</sup>Ct value for the Xpert Flu result = 37.8. Discrepancy testing with the Liat Flu assay was positive (agreement with Xpert Flu).

## DISCUSSION

The results here show PPA for the MAX SARS-CoV-2/Flu assay with reference assays meet FDA-EUA acceptance criteria for detection of SARS-CoV-2 (96.2%), Flu A (100%; with a lower bound 95%CI of 93.9%), and Flu B (98.3%; with a lower bound 95%CI of 91.1%). Similarly, compared to reference methods, the MAX SARS-CoV-2/Flu assay was associated with NPA values for detection of SARS-CoV-2 (100%), Flu A (98.9%; with a lower bound 95%CI of 94.0%), and Flu B (100%; with a lower bound 95%CI of 95.9%) that all met FDA acceptance criteria. Discordant results were further tested with the third assays, BioFire SARS-CoV-2 assay SARS-CoV-2 and Liat Flu assay for Flu A/B. All discrepant results were associated with high Ct values. Thus, with its high PPA and NPA for SARS-CoV-2, Flu A, and Flu B, this multiplex assay should reduce specimen collection time and the amount of supplies and reagents necessary to test for both COVID-19 and Flu.

Different approaches are currently available for the detection of SARS-CoV-2 and Flu A/B for the diagnosis of both COVID-19 and influenza, respectively.(12, 17) Although culture-based assays were originally utilized to establish an influenza diagnosis, RT-PCR-based technology for diagnosis of influenza currently represents the laboratory method of choice due to its relatively high analytic and clinical sensitivity, as well as short turn-around time.(18) Likewise, RT-PCR-based assays appear to have higher sensitivity for detection for SARS-CoV-2 compared to culture-based assays.(19) Rapid testing, such as immunochromatic techniques are used to detect viral antigen, have been developed for detection of both SARS-CoV-2 and Flu A/B.(11, 12) Although rapid tests carry advantages, such as decreased time-to-result and ease of implementation in decentralized health care settings, RT-PCR-based assays have increased analytical sensitivity compared to rapid tests.(12) Ultimately, multiple factors should be considered before determining which strategy should be employed. For example, hospitals and their associated laboratory partners, which have established a streamlined workflow and a relatively fast turn-around-time, can effectively employ RT-PCR-based assays—especially for patients admitted and managed according to their symptoms. This strategy carries the benefit of high sensitivity and the ability to rule out etiologic agents with a high degree of assurance.

The MAX SARS-CoV-2/Flu assay utilizes the same multiplexed primers and probes targeting RNA from the nucleocapsid phosphoprotein gene (N1 and N2 regions) of the SARS-CoV-2 virus as shown in the previous FDA-EUA approved MAX SARS-CoV-2 assay.(20) The MAX SARS-CoV-2/Flu assay also includes additional primers and probes recognizing a conserved region of the matrix protein M1 gene for influenza A and conserved regions of the matrix protein M1 gene and hemagglutinin (HA) gene for influenza B.(21) A positive result for SARS-CoV-2 with a low Ct value may be indicative of active infection, however, this result does not rule out bacterial infection or co-infection with other viruses.(20) This is important in the case of SARS-CoV-2, as asymptomatic infections are possible and the positive results require clinical judgement. While the MAX SARS-CoV-2/Flu assay can detect SARS-CoV-2 and influenza A and B virus, it is not intended to detect influenza C virus.(21) The clinical presentation and contact history of an individual along with other diagnostic information is necessary to determine the actual infection status.

If the coincidence of high rates of both COVID-19 and influenza cases occurs during a respiratory virus season, differential diagnosis for the appropriate therapeutic approach could be challenging. Although COVID-19 and influenza spread through a similar transmission mechanism and have overlapping symptomology, specific differences between the diseases do exist. For example, COVID-19 seems to involve a longer time to symptom onset than influenza, and may cause more severe illness in vulnerable populations once symptoms develop.(22) Also, the therapeutic approach for COVID-19 patients and influenza patients is not similar. While the impact of anti-viral drug for influenza patients is standardized, this is not the case for the treatment SARS-CoV-2-positive individuals. Medications such as remdesivir and/or corticosteroids have been used to treat COVID-19 patients, however, these two approaches are not approved for use in influenza patients.(6) Distinguishing the diagnosis of COVID-19 and influenza, therefore, will be an important component for proper patient triage, management, and treatment.

### Limitations

This research was conducted by using materials obtained from pre-selected frozen remnants, received after routine care. A study involving prospective collection would better inform on the positive and negative predictive values of the assay.

### Conclusions

The MAX SARS-CoV-2/Flu assays met US FDA-EUA acceptance criteria for SARS-CoV-2 and Flu A/B detection. Dual detection of the etiologic agents causing COVID-19 and influenza will allow differentiation for those exhibiting common symptoms between the two diseases. This assay should help optimize patient management by decreasing the time and resources required for dual testing. Ultimately, the dual detection method should facilitate an informed decision by physicians on the appropriate treatment for patients exhibiting similar symptoms between the two diseases.

## Data Availability

All data are available upon request.

## ACKNOWLEDGEMENTS

We thank Karen Eckert and Karen Yanson (Becton, Dickinson and Company, BD Life Sciences – Integrated Diagnostic Solutions) for their input on the study logistics and content of this manuscript and editorial assistance. We also thank Yu-Chih Lin and Devin Gary for the editorial assistance. We thank Stanley Chao, Aojun Li, and Yongqiang Zhang (Becton, Dickinson and Company, BD Life Sciences – Integrated Diagnostic Solutions) for statistical support. The individuals acknowledged here have no additional funding or additional compensation to disclose. We are grateful to the study participants who allowed this work to be performed.

## AUTHOR CONTRIBUTIONS

**Sonia Paradis:** Conceptualization, Resources, Writing – Original Draft, Writing – Review & Editing, Visualization, Project administration. **Elizabeth Lockamy:** Methodology, Formal analysis, Investigation, Resources, Data curation, Writing – Review & Editing. **Charles K. Cooper:** Conceptualization, Supervision, Writing – Review & Editing. **Stephen Young:** Conceptualization, Methodology, Formal analysis, Investigation, Resources, Data Curation, Writing – Review & Editing, Supervision, Project administration, Funding acquisition.

## FUNDING

This study was funded by Becton, Dickinson and Company; BD Life Sciences—Integrated Diagnostics Solutions. Non-BD employee authors received research funds to support their work for this study.

## POTENTIAL CONFLICTS OF INTEREST

The authors disclose the following conflicts of interest: SP, EL, and CKC are employees of Becton, Dickinson and Company; SY, None.

---

**Table S1.** Study assays and locations for assay performance.

| **Assay** | **BD (Sparks, MD)** | **TriCore (Albuquerque, NM)** |
| --- | --- | --- |
| BD MAX SARS-CoV-2/Flu<sup>a,b</sup> | √ |  |
| BioGx SARS-CoV-2<sup>a, c</sup> | √ |  |
| Xpert Flu<sup>b, d</sup> |  | √ |
| BioFire SARS-CoV-2<sup>a, e</sup> | √ |  |
| Liat Flu<sup>b, f</sup> |  | √ |

<sup>a</sup>Specimens for SARS-CoV-2 testing were obtained from New York Biologics (Southampton, NY) and Trans-Hit Bio (Laval, QC, Canada).
<sup>b</sup>Specimens for Flu A/B testing were obtained from New York Biologics.
<sup>c</sup>Utilized for reference test during detection of SARS-CoV-2.
<sup>d</sup>Utilized for reference test during detection of Flu A/B.
<sup>e</sup>Utilized for discordant test involving detection of SARS-CoV-2.
<sup>f</sup>Utilized for discordant test involving detection of Flu A/B.

## Notes

### Competing Interest Statement

The authors have declared no competing interest.

### Clinical Trial

This research was conducted by using materials obtained from pre-selected frozen remnants, received after routine care.

### Author Declarations

New York Biologics holds two IRB approvals for residual specimen collection: one from Western IRB (WIRB) and the other from Ethical & Independent Review Services (EIRS). Trans-Hit Bio partners with academic and hospital-based biobanks and other clinical collection sites. These samples were collected under their biobank umbrella protocol (approved by the Valleywise Health Institutional Review Board) that allows for the collection of various biospecimens including NP swabs. Under this protocol, ICF is waived for diagnostic remnants (as was the case with the NP swabs)

